# Diagnostic signature for Heart Failure with Preserved Ejection Fraction (HFpEF): A Machine Learning Approach Using Multi-Modality Electronic Health Record Data

**DOI:** 10.1101/2021.11.18.21266560

**Authors:** Nazli Farajidavar, Kevin O’Gallagher, Daniel Bean, Adam Nabeebaccus, Rosita Zakeri, Daniel Bromage, Zeljko Kraljevic, James TH Teo, Richard J Dobson, Ajay M Shah

## Abstract

**Aims:** Heart failure with preserved ejection fraction (HFpEF) is thought to be highly prevalent yet remains underdiagnosed. We sought to develop a data-driven diagnostic model to predict from electronic health records (EHR) the likelihood of HFpEF among patients with unexplained dyspnea and preserved left ventricular EF.

**Methods & Results:** The derivation cohort comprised patients with dyspnea and echocardiography results. Structured and unstructured data were extracted using an automated informatics pipeline. Patients were retrospectively diagnosed as HFpEF (cases), non-HF (control cohort I), or HF with reduced EF (HFrEF; control cohort II). The ability of clinical parameters and investigations to discriminate cases from controls was evaluated by extreme gradient boosting. A likelihood scoring system was developed and validated in a separate test cohort.

The derivation cohort included 1585 consecutive patients: 133 cases of HFpEF (9%), 194 non-HF cases (Control cohort I) and 1258 HFrEF cases (Control cohort II). Two HFpEF diagnostic signatures were derived, comprising symptoms, diagnoses and investigation results. A final prediction model was generated based on the averaged likelihood scores from these two models. In a validation cohort consisting of 269 consecutive patients (with 66 HFpEF cases (24.5%)), the diagnostic power of detecting HFpEF had an AUROC of 90% (P<0.001) and average precision (AP) of 74%.

**Conclusion:** This diagnostic signature enables discrimination of HFpEF from non-cardiac dyspnea or HFrEF from EHR and can assist in the diagnostic evaluation in patients with unexplained dyspnea.

## INTRODUCTION

Heart Failure with preserved ejection fraction (HFpEF) is a highly prevalent yet under-diagnosed clinical syndrome[1, 2]. The hallmarks are the signs and symptoms of heart failure (HF) and a preserved left ventricular ejection fraction (LVEF). HFpEF is thought to be underpinned by structural and functional abnormalities of both the heart and vasculature. Patients with HFpEF typically display diastolic dysfunction[3, 4] and other abnormalities such as vascular stiffening[5] and impaired ventricular-vascular coupling[6-10]. Unlike HF with reduced Ejection Fraction (HFrEF), no evidence-based therapies are available for HFpEF[11-13]. This may in part reflect the heterogeneity of HFpEF pathophysiology as well as issues of clinical trial design[13-15].

While the diagnosis of HFpEF is straightforward in acutely decompensated patients, stable euvolemic patients present a greater challenge[16]. Exertional dyspnea and fatigue are non-specific symptoms that occur in many other conditions, including obesity and physical deconditioning. Expert transthoracic echocardiography (ideally with exercise) or invasive cardiac catheterization to document raised LV filling pressures may not be immediately available to the non-specialist. A recent study found that among more than 44,000 community-based patients likely to have HF, only 50% had a documented LVEF[17]. Furthermore, those eventually diagnosed as having HFpEF required many more pre-diagnosis investigations and consultations than HFrEF patients.

In previous epidemiological studies, identification and extraction of HFpEF cases from Electronic Health Records (EHR) has typically relied on diagnostic codes, additional medical record abstraction, and/or adjudication based on various expert criteria e.g. European Society of Cardiology criteria[18]. The EHR is however increasingly amenable to rapid and automated extraction of multiple clinical parameters, including the use of advanced natural language processing (NLP) algorithms to identify clinical concepts recorded in the unstructured text[19-21].

The aim of this study was to extract and analyze data from the EHR to develop an automated approach to identify patients likely to have HFpEF.

## METHODS

### Approvals

This project was conducted under London South East Research Ethics Committee approval (reference 18/LO/2048) granted to the King’s Electronic Records Research Interface (KERRI), project ID 202020201.

### Derivation Cohort

We performed a retrospective study using de-identified data of patients attending King’s College Hospital NHS Foundation Trust (KCH) in London (UK) between 2000 and 2019. We focused on patients who had undergone echocardiography as part of their inpatient or outpatient evaluation. With this starting point, a number of different patient cohorts were derived based on the LVEF, confirmed or possible HF, symptoms of dyspnea, and NT-proBNP (or BNP) level (see **Supplementary materials Sections I and II**). We identified confirmed HFpEF cases and two control cohorts: those with no evidence of HF (non-HF, Control cohort I) and those with HFrEF (Control cohort II). HFpEF cases were defined as patients with a preserved LVEF ≥50% (with no evidence of LVEF <50% at any stage), a confirmed diagnosis of HF based on discharge ICD10 codes I50.0, I50.1 or I50.9, dyspnea, and a raised NT-proBNP or BNP level (according to age-specific thresholds), in accordance with ESC diagnostic criteria[18]. Patients with valvular heart disease (ICD10 codes I05-I09 and I35) were excluded.

### Test Cohorts

We generated 4 test cohorts from patients who lacked at least one of the above diagnostic features for a confirmed diagnosis of HFpEF (see **Supplementary Table S1 and Flowchart S1**). We randomly sampled 100 patients from each of these four test subsets for analysis and removed samples where the clinical annotations disagreed or there was more than 70% missingness in signature predictors, leaving 269 in total.

### Data extraction and evaluation

Clinical and demographic data were retrieved from the structured and unstructured components of the EHR using the CogStack informatics platform[20]. Automated parsing of the EHR was achieved with a state-of-the-art enterprise search and well-validated natural language processing (NLP) tools, including MedCAT[22] and the Unified Medical Language System repository[23] as previously used by our group.[24] Clinical term extraction was restricted to concepts which represent clinical findings, diseases (apart from HF), medications, and signs and symptoms. This was linked to searches of structured data from an internal database containing echocardiographic data and ICD codes. Continuous variables were cleaned prior to cohort selection; e.g. conversion of text references of LVEF to numerical values and removal of measurement outliers (see **Supplementary material Section III**). We used both platforms to arbitrate discrepancies in our derivation dataset as neither source proved to be comprehensive, in line with previous work[20, 21].

Echocardiographic data were based on studies performed according to British Society of Echocardiography guidelines[25] (which are consistent with American and European guidelines)[26]. Structured data recorded in echocardiography results were boosted with numerical data reported in the EHR text. Additionally, when appropriate (e.g. patient had echocardiography but a numerical value for LVEF was not documented) we used a deep learning model to infer whether the LVEF was preserved based on the echocardiography report (see **Supplementary materials Section III**).

BNP or NT-proBNP results were obtained from samples drawn at any time in the study period and the maximum value for each subject was used.

All cases in the derivation dataset that were identified by the data pipeline as HFpEF were validated by manual review of the EHR by a cardiologist.

### Potential modeling predictors

A binary diagnostic outcome indicating the presence or absence of HFpEF was considered for modeling. Potential predictors to be included in a diagnostic signature included those used in previous HFpEF epidemiological studies[14, 15]. In addition, we adopted a comprehensive approach that included physiological variables, laboratory results, echocardiographic data and clinical concept references[27]. Structured data were collected within a two-month temporal window around the last echocardiography result (or NTproBNP/BNP test result if available). Unstructured data were analyzed from the entire EHR prior to the date of the echocardiography result for each patient.

We made a second level predictor grouping according to whether the variables were initially recorded as (a) structured data: demographic and physiological parameters, and laboratory and echocardiography measurements; or (b) unstructured text in the EHR, extracted via the NLP platform. We adopted the bag-of-words[28] approach to transform clinical concept annotation into word vectors for modeling purposes. Concepts which were mentioned in <10% of the derivation cohort were excluded. Data from the other predictor categories were collected and imputed prior to training, using the k-nearest neighbor (Scikit-learn python package v0.22) after min-max normalization. Following imputation, data items were rescaled into their original range to preserve the explainability of the final model.

### Data modelling, feature selection and validation

We used the tree-based multivariable extreme gradient boosting[29] algorithm (XGBoost, python package v0.9) for modeling, enabling inclusion of mixed data types and smooth handling of missing values and sparsity issues. As such, when a value is missing in the sparse predictor vector, the instance is classified into a default direction (see[29] for further details) that is learnt as optimal using derivation data.

SHAP[30] analysis (SHapley Additive exPlanations; SHAP python package v0.33) was used to order the predictors according to their prominence in discriminating cases from controls. Once the full model was created, we took a stepwise forward insertion scheme to include the more significant variables one at a time, in order to determine the minimal number of predictors that gave an acceptable performance relative to the use of all predictors. The final predictive models were trained and evaluated using the obtained optimal subset of predictors.

Model validation was undertaken in the test cohorts described earlier, using clinical assessment criteria from the H_2_FPEF score[16] as a comparator. A random sample of 400 patients from the test datasets was manually reviewed by two teams each comprising two cardiologists, in order to validate diagnoses. Any cases of clinician disagreement were removed from the evaluation, leaving a total of 269 patients in the test datasets (see Results, **Table 1**).

**Table 1.**
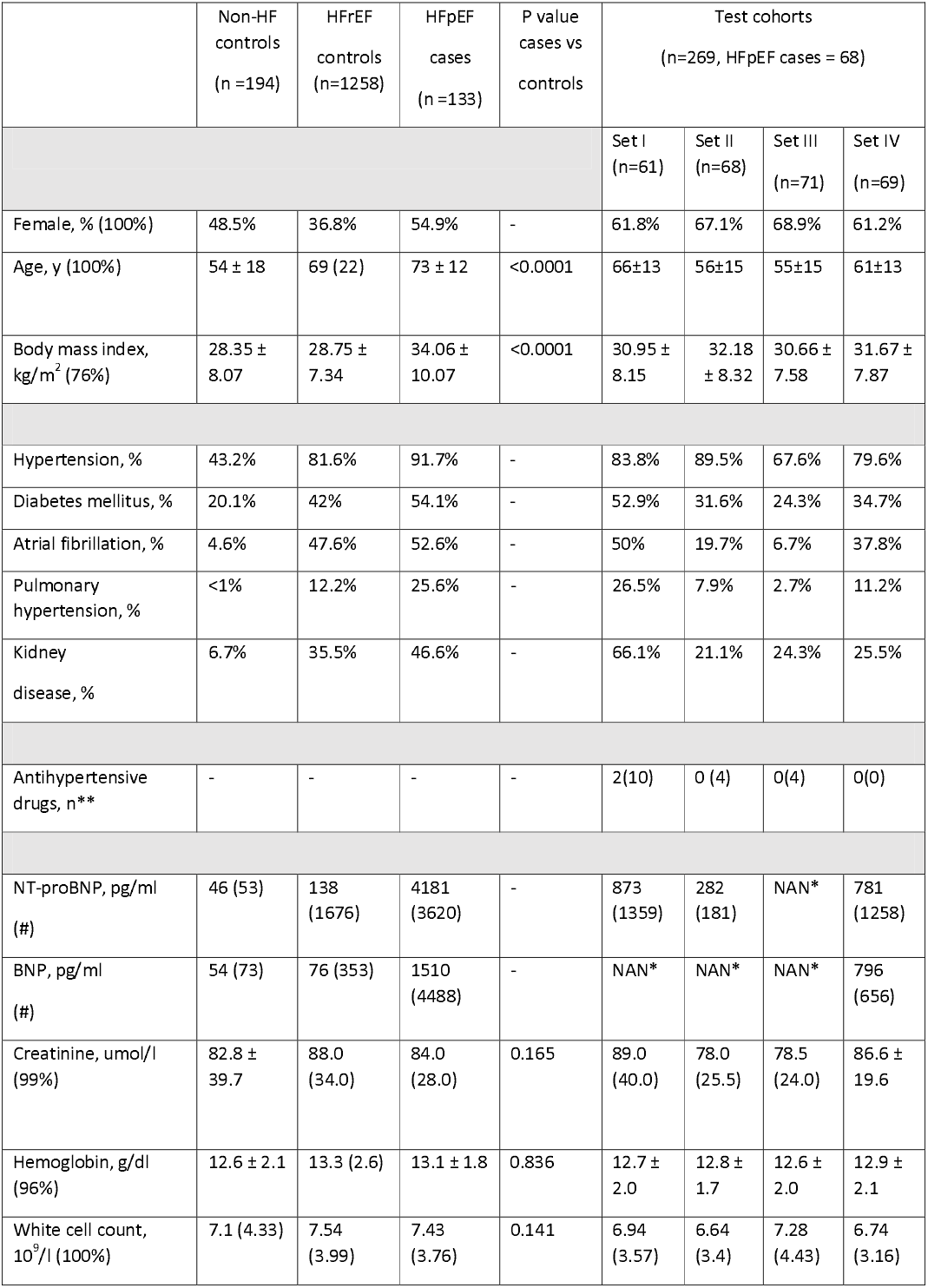
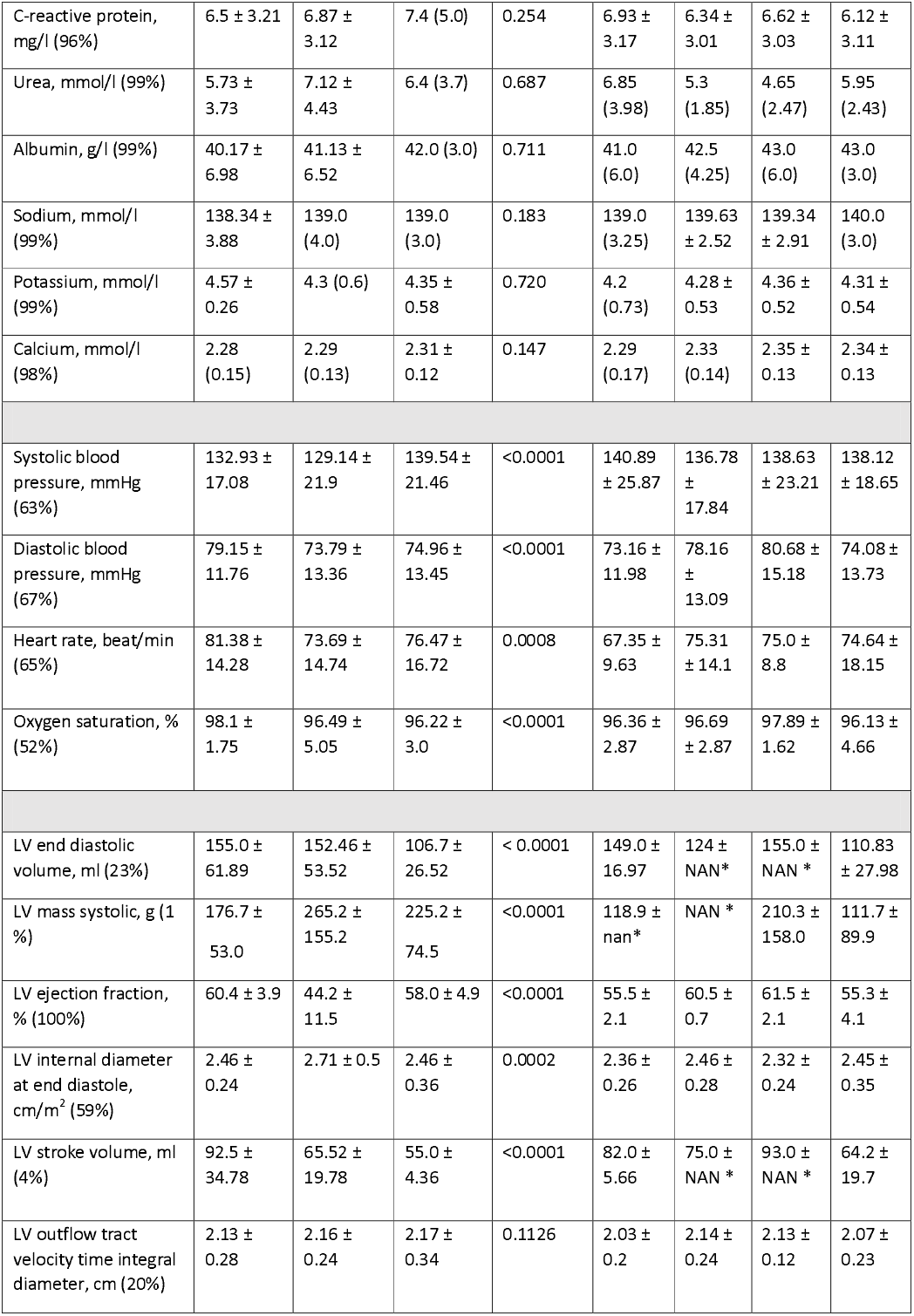
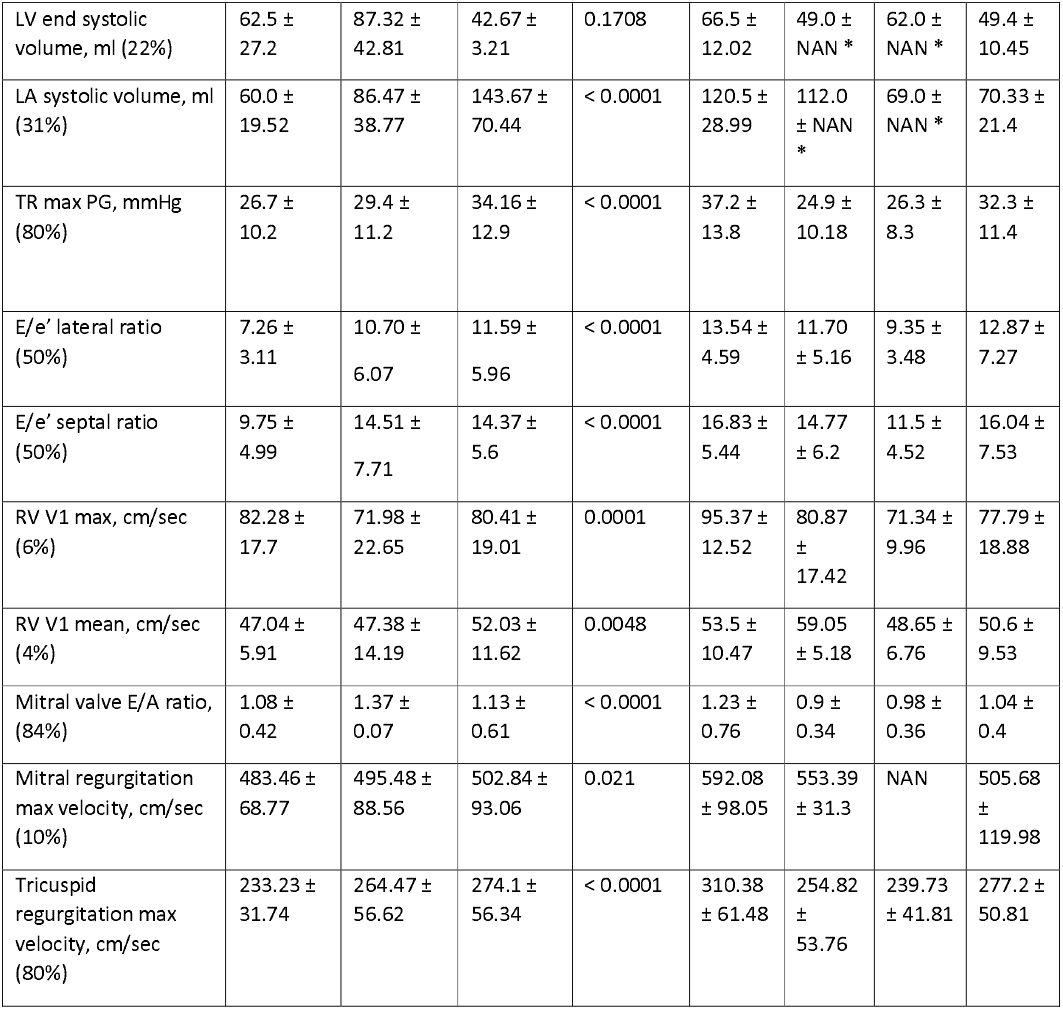
Baseline characteristics of patients. The mean and SD (standard deviation) were obtained where the predictor distribution follows a normal distribution, whereas for predictors with a skewed distribution, the median and interquartile range (25^th^-75^th^) were used to report the statistics. To evaluate the distributional differences between cases and controls, the Mann-Whitney U test or the t test was acquired, where appropriate. Values in parentheses next to each predictor name indicate the data availability percentage. * Constraint-free assumption on our test sets resulted in predictors with either a singular value or a high proportion of missing values. In such cases, the computation of common statistics was not pragmatic and hence the NAN (Not A Number) value was reported, instead. ** This predictor is only computed in the test cohort to enable the comparison with the H_2_FPEF score. # 92.45% of HFpEF cases and controls had a BNP or pro-BNP level available. Set I: patients with normal EF, no/normal BNP record, a HF ICD10 code and at least one HF and dyspnea reference in their EHR. Set II: patients with normal EF, no/normal BNP record, no HF diagnostic code and at least one HF and dyspnea reference in their EHR. Set III: patients with normal EF, no BNP record, no HF diagnostic code nor HF reference in the EHR, at least one report of their dyspnea in their EHR. Set IV: patients with normal EF, raised BNP result with HF and dyspnea reference in their EHR but no HF diagnosis documented (HF: heart failure, EF: ejection fraction, rEF: reduced EF, BNP: brain-natriuretic peptide test, EHR: electronic health record). The following ICD10 codes were used to define the comorbidities: Hypertension: I10-I15, I60-I69; Diabetes mellitus: E10-E14; Atrial fibrillation: I48; Pulmonary hypertension: I27; Kidney Disease: N18, N28, I12-I15

|  | Non-HF<br>controls<br>(n =194) | HFrEF<br>controls<br>(n=1258) | HFpEF<br>cases<br>(n =133) | P value<br>cases vs<br>controls | Test cohorts<br>(n=269, HFpEF cases = 68) |  |  |  |
| --- | --- | --- | --- | --- | --- | --- | --- | --- |
|  |  |  |  |  | Set I<br>(n=61) | Set II<br>(n=68) | Set III<br>(n=71) | Set IV<br>(n=69) |
| Female, % (100%) | 48.5% | 36.8% | 54.9% | - | 61.8% | 67.1% | 68.9% | 61.2% |
| Age, y (100%) | 54 ± 18 | 69 (22) | 73 ± 12 | <0.0001 | 66±13 | 56±15 | 55±15 | 61±13 |
| Body mass index,<br>kg/m<sup>2</sup> (76%) | 28.35 ±<br>8.07 | 28.75 ±<br>7.34 | 34.06 ±<br>10.07 | <0.0001 | 30.95 ±<br>8.15 | 32.18<br>± 8.32 | 30.66 ±<br>7.58 | 31.67 ±<br>7.87 |
| Hypertension, % | 43.2% | 81.6% | 91.7% | - | 83.8% | 89.5% | 67.6% | 79.6% |
| Diabetes mellitus, % | 20.1% | 42% | 54.1% | - | 52.9% | 31.6% | 24.3% | 34.7% |
| Atrial fibrillation, % | 4.6% | 47.6% | 52.6% | - | 50% | 19.7% | 6.7% | 37.8% |
| Pulmonary<br>hypertension, % | <1% | 12.2% | 25.6% | - | 26.5% | 7.9% | 2.7% | 11.2% |
| Kidney<br>disease, % | 6.7% | 35.5% | 46.6% | - | 66.1% | 21.1% | 24.3% | 25.5% |
| Antihypertensive<br>drugs, n** | - | - | - | - | 2(10) | 0 (4) | 0(4) | 0(0) |
| NT-proBNP, pg/ml<br>(#) | 46 (53) | 138<br>(1676) | 4181<br>(3620) | - | 873<br>(1359) | 282<br>(181) | NAN* | 781<br>(1258) |
| BNP, pg/ml<br>(#) | 54 (73) | 76 (353) | 1510<br>(4488) | - | NAN* | NAN* | NAN* | 796<br>(656) |
| Creatinine, umol/l<br>(99%) | 82.8 ±<br>39.7 | 88.0<br>(34.0) | 84.0<br>(28.0) | 0.165 | 89.0<br>(40.0) | 78.0<br>(25.5) | 78.5<br>(24.0) | 86.6 ±<br>19.6 |
| Hemoglobin, g/dl<br>(96%) | 12.6 ± 2.1 | 13.3 (2.6) | 13.1 ± 1.8 | 0.836 | 12.7 ±<br>2.0 | 12.8 ±<br>1.7 | 12.6 ±<br>2.0 | 12.9 ±<br>2.1 |
| White cell count,<br>10<sup>9</sup>/l (100%) | 7.1 (4.33) | 7.54<br>(3.99) | 7.43<br>(3.76) | 0.141 | 6.94<br>(3.57) | 6.64<br>(3.4) | 7.28<br>(4.43) | 6.74<br>(3.16) |
| C-reactive protein, mg/l (96%) | 6.5 ± 3.21 | 6.87 ± 3.12 | 7.4 (5.0) | 0.254 | 6.93 ± 3.17 | 6.34 ± 3.01 | 6.62 ± 3.03 | 6.12 ± 3.11 |
| Urea, mmol/l (99%) | 5.73 ± 3.73 | 7.12 ± 4.43 | 6.4 (3.7) | 0.687 | 6.85 (3.98) | 5.3 (1.85) | 4.65 (2.47) | 5.95 (2.43) |
| Albumin, g/l (99%) | 40.17 ± 6.98 | 41.13 ± 6.52 | 42.0 (3.0) | 0.711 | 41.0 (6.0) | 42.5 (4.25) | 43.0 (6.0) | 43.0 (3.0) |
| Sodium, mmol/l (99%) | 138.34 ± 3.88 | 139.0 (4.0) | 139.0 (3.0) | 0.183 | 139.0 (3.25) | 139.63 ± 2.52 | 139.34 ± 2.91 | 140.0 (3.0) |
| Potassium, mmol/l (99%) | 4.57 ± 0.26 | 4.3 (0.6) | 4.35 ± 0.58 | 0.720 | 4.2 (0.73) | 4.28 ± 0.53 | 4.36 ± 0.52 | 4.31 ± 0.54 |
| Calcium, mmol/l (98%) | 2.28 (0.15) | 2.29 (0.13) | 2.31 ± 0.12 | 0.147 | 2.29 (0.17) | 2.33 (0.14) | 2.35 ± 0.13 | 2.34 ± 0.13 |
| Systolic blood pressure, mmHg (63%) | 132.93 ± 17.08 | 129.14 ± 21.9 | 139.54 ± 21.46 | <0.0001 | 140.89 ± 25.87 | 136.78 ± 17.84 | 138.63 ± 23.21 | 138.12 ± 18.65 |
| Diastolic blood pressure, mmHg (67%) | 79.15 ± 11.76 | 73.79 ± 13.36 | 74.96 ± 13.45 | <0.0001 | 73.16 ± 11.98 | 78.16 ± 13.09 | 80.68 ± 15.18 | 74.08 ± 13.73 |
| Heart rate, beat/min (65%) | 81.38 ± 14.28 | 73.69 ± 14.74 | 76.47 ± 16.72 | 0.0008 | 67.35 ± 9.63 | 75.31 ± 14.1 | 75.0 ± 8.8 | 74.64 ± 18.15 |
| Oxygen saturation, % (52%) | 98.1 ± 1.75 | 96.49 ± 5.05 | 96.22 ± 3.0 | <0.0001 | 96.36 ± 2.87 | 96.69 ± 2.87 | 97.89 ± 1.62 | 96.13 ± 4.66 |
| LV end diastolic volume, ml (23%) | 155.0 ± 61.89 | 152.46 ± 53.52 | 106.7 ± 26.52 | < 0.0001 | 149.0 ± 16.97 | 124 ± NAN* | 155.0 ± NAN * | 110.83 ± 27.98 |
| LV mass systolic, g (1 %) | 176.7 ± 53.0 | 265.2 ± 155.2 | 225.2 ± 74.5 | <0.0001 | 118.9 ± nan* | NAN * | 210.3 ± 158.0 | 111.7 ± 89.9 |
| LV ejection fraction, % (100%) | 60.4 ± 3.9 | 44.2 ± 11.5 | 58.0 ± 4.9 | <0.0001 | 55.5 ± 2.1 | 60.5 ± 0.7 | 61.5 ± 2.1 | 55.3 ± 4.1 |
| LV internal diameter at end diastole, cm/m<sup>2</sup> (59%) | 2.46 ± 0.24 | 2.71 ± 0.5 | 2.46 ± 0.36 | 0.0002 | 2.36 ± 0.26 | 2.46 ± 0.28 | 2.32 ± 0.24 | 2.45 ± 0.35 |
| LV stroke volume, ml (4%) | 92.5 ± 34.78 | 65.52 ± 19.78 | 55.0 ± 4.36 | <0.0001 | 82.0 ± 5.66 | 75.0 ± NAN * | 93.0 ± NAN * | 64.2 ± 19.7 |
| LV outflow tract velocity time integral diameter, cm (20%) | 2.13 ± 0.28 | 2.16 ± 0.24 | 2.17 ± 0.34 | 0.1126 | 2.03 ± 0.2 | 2.14 ± 0.24 | 2.13 ± 0.12 | 2.07 ± 0.23 |
| LV end systolic volume, ml (22%) | 62.5 ± 27.2 | 87.32 ± 42.81 | 42.67 ± 3.21 | 0.1708 | 66.5 ± 12.02 | 49.0 ± NAN * | 62.0 ± NAN * | 49.4 ± 10.45 |
| LA systolic volume, ml (31%) | 60.0 ± 19.52 | 86.47 ± 38.77 | 143.67 ± 70.44 | < 0.0001 | 120.5 ± 28.99 | 112.0 ± NAN * | 69.0 ± NAN * | 70.33 ± 21.4 |
| TR max PG, mmHg (80%) | 26.7 ± 10.2 | 29.4 ± 11.2 | 34.16 ± 12.9 | < 0.0001 | 37.2 ± 13.8 | 24.9 ± 10.18 | 26.3 ± 8.3 | 32.3 ± 11.4 |
| E/e' lateral ratio (50%) | 7.26 ± 3.11 | 10.70 ± 6.07 | 11.59 ± 5.96 | < 0.0001 | 13.54 ± 4.59 | 11.70 ± 5.16 | 9.35 ± 3.48 | 12.87 ± 7.27 |
| E/e' septal ratio (50%) | 9.75 ± 4.99 | 14.51 ± 7.71 | 14.37 ± 5.6 | < 0.0001 | 16.83 ± 5.44 | 14.77 ± 6.2 | 11.5 ± 4.52 | 16.04 ± 7.53 |
| RV V1 max, cm/sec (6%) | 82.28 ± 17.7 | 71.98 ± 22.65 | 80.41 ± 19.01 | 0.0001 | 95.37 ± 12.52 | 80.87 ± 17.42 | 71.34 ± 9.96 | 77.79 ± 18.88 |
| RV V1 mean, cm/sec (4%) | 47.04 ± 5.91 | 47.38 ± 14.19 | 52.03 ± 11.62 | 0.0048 | 53.5 ± 10.47 | 59.05 ± 5.18 | 48.65 ± 6.76 | 50.6 ± 9.53 |
| Mitral valve E/A ratio, (84%) | 1.08 ± 0.42 | 1.37 ± 0.07 | 1.13 ± 0.61 | < 0.0001 | 1.23 ± 0.76 | 0.9 ± 0.34 | 0.98 ± 0.36 | 1.04 ± 0.4 |
| Mitral regurgitation max velocity, cm/sec (10%) | 483.46 ± 68.77 | 495.48 ± 88.56 | 502.84 ± 93.06 | 0.021 | 592.08 ± 98.05 | 553.39 ± 31.3 | NAN | 505.68 ± 119.98 |
| Tricuspid regurgitation max velocity, cm/sec (80%) | 233.23 ± 31.74 | 264.47 ± 56.62 | 274.1 ± 56.34 | < 0.0001 | 310.38 ± 61.48 | 254.82 ± 53.76 | 239.73 ± 41.81 | 277.2 ± 50.81 |

### Statistical analysis of predictors

Data are presented as mean and standard deviation (SD) or median and interquartile range (IQR) as appropriate. Differences between cases and controls were evaluated by the Mann-Whitney U test or unpaired t test, as appropriate. The area under the receiver-operating characteristic curve (AUROC), F1-score (macro and weighted^2^) and average precision (AP) were used as performance metrics.

A stratified 5-fold cross-validation scheme (to ensure each fold is a good representative of the whole data in terms of class prevalence) was utilized for feature selection and derivation set validation. As such, the derivation data was divided into five subsets, four of which were used for training the model and the final one for validation/testing. The derivation and test subsets were shuffled until all five subsets were evaluated. The final performance was then reported as mean and standard deviation of all five tests.

The AUROC and AP were used as performance metrics and the Kappa statistic was used to measure the inter-rater agreement of proposed models. All tests were 2-sided, with P<0.05 considered significant.

To evaluate the generalizability of the model to a new sample, Harrell optimism was calculated with 1000 boot-strap replicates[31]. To evaluate discrimination power of the proposed model beyond existing criteria, we compared the model’s AUROCs and AP performance against the recently proposed H_2_FPEF scoring system[16] using the Random Forest (predecessor to XGBoost).

Statistical analyses were performed in Python 3 using SciPy and Scikit-learn packages (v0.22).

### Data availability

The data included in the study will not be made available to other researchers due to hospital information governance regulations. However, we will share our models and the analytical methods to facilitate the replication of the study on data collected from other hospitals.

## RESULTS

1854 patients were included in the study of whom 1585 were in the derivation cohort (**Table 1**). HFpEF patients in the derivation cohort (n=133) were older than those with non-HF or HFrEF, with a higher proportion of females and a higher BMI. They also had a higher prevalence of hypertension, atrial fibrillation, diabetes and chronic kidney disease. Systolic and diastolic pressures were higher in the HFpEF group compared to HFrEF. Patients with HFpEF had lower end-diastolic and end-systolic volumes and higher septal E/e’ ratios than the non-HF control group.

### Structured, unstructured and combined signatures for HFpEF diagnosis

We initially divided the predictors into two sets based on the source of data being structured data or clinical concepts and conditions extracted from the unstructured historical EHR (*see Methods*). We excluded the BNP/NT-proBNP assessment data and HF concept references from both predictor sets to avoid biasing models by information on outcome. Separate XGBoost models were trained on each predictor set. SHAP analysis was adopted to select the optimal number of features from each predictor set using five-fold cross-validation. We then compared the discriminant power of these signatures to distinguish HFpEF cases either from non-HF patients (Control set I) or HFrEF patients (Control set II).

The minimum number of variables required to maintain an acceptable level of performance for each model were selected (**Figure 1**). Following an early-fusion modeling strategy, we merged the selected predictors from the two sets of structured and unstructured variables and trained an XGBoost model for discrimination and termed the derived signature as the combined signature.

**Figure 1.**
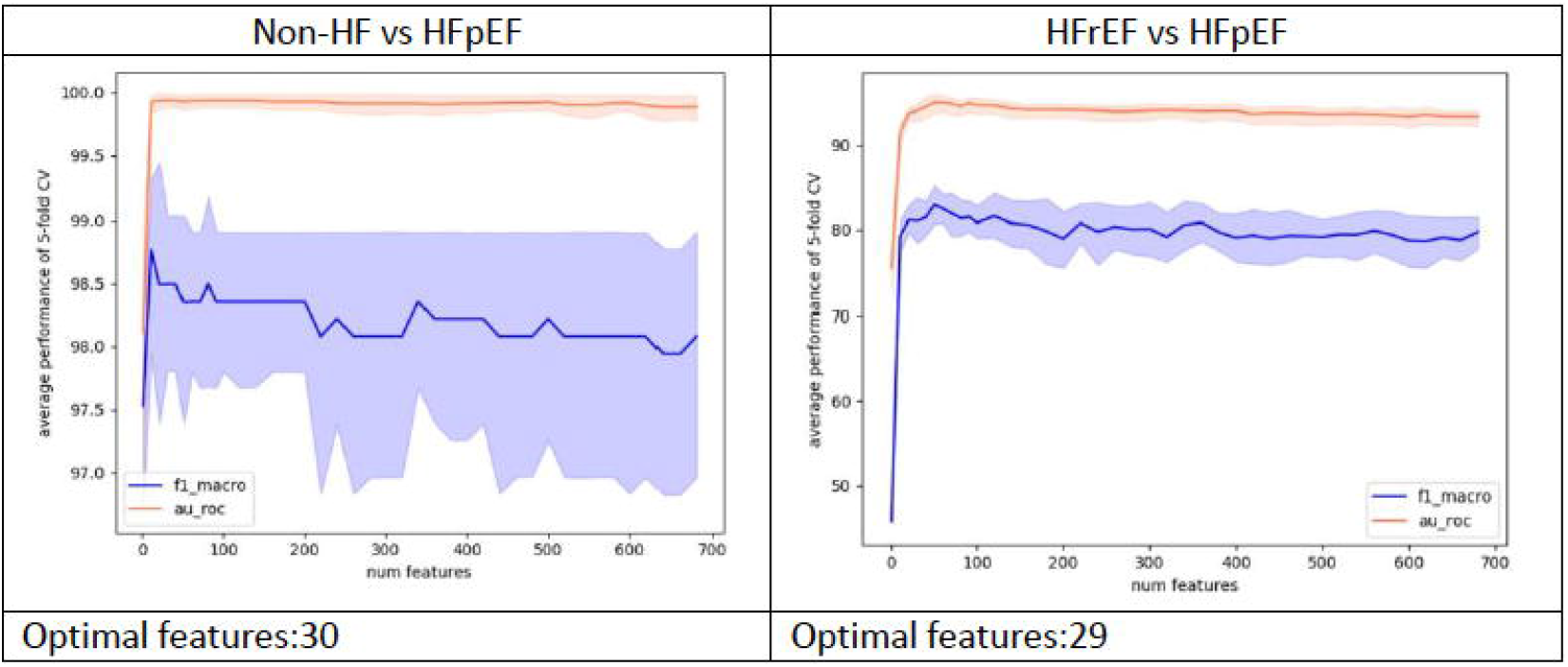
Feature selection analysis. Features were incrementally utilized for training the models to ensure a performance within ±2 units of the AUROC and f1-macro metrics in 5-fold cross-validation setup. Blue: f1-macro, Red: AUROC

SHAP analysis to assess feature importance showed that individual predictors had different value in discrimination of HFpEF versus non-HF or HFrEF (**Figure 2**). For example, dyspnea and pharmacologic substance were the most prominent predictors in discrimination against non-HF whereas EF was most important for discrimination against HFrEF. However, many of the features (e.g. age, patient address) were common to the two groups. The text references to “patient address” and “pharmacologic substance” were surrogate predictors of the number of complete hospital admissions. **(Figure 2)**.

**Figure 2.**
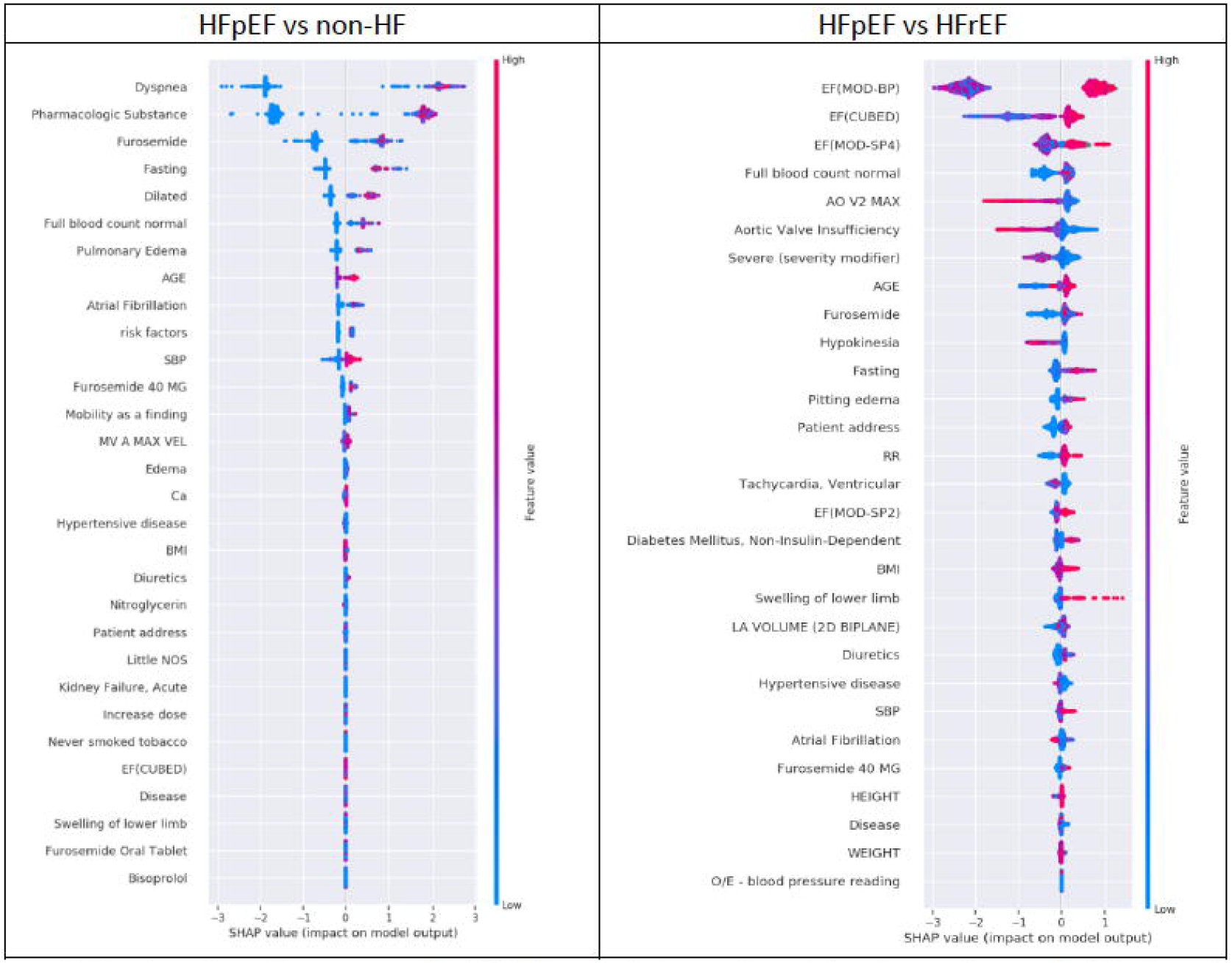
Feature importance using SHAP analysis in combined signatures. Denser distribution of red points at the positive quadrant of the plot is representative of higher values of a given predictor’s contribution in characterizing the positive class distribution i.e. in characterizing HFpEF.

The combined signature model for discrimination of HFpEF from HFrEF showed an enhanced AUROC performance and F1-measure score as compared to the single-view models in the 5-fold cross-validation evaluation in our developmental dataset (**Table 2**). The performance enhancement of the combined model in discriminating HFpEF from non-HF was less significant. This was due to dominancy of the unstructured predictors in this combined signature (see **Figure 2** and **Table 3**).

**Table 2.**
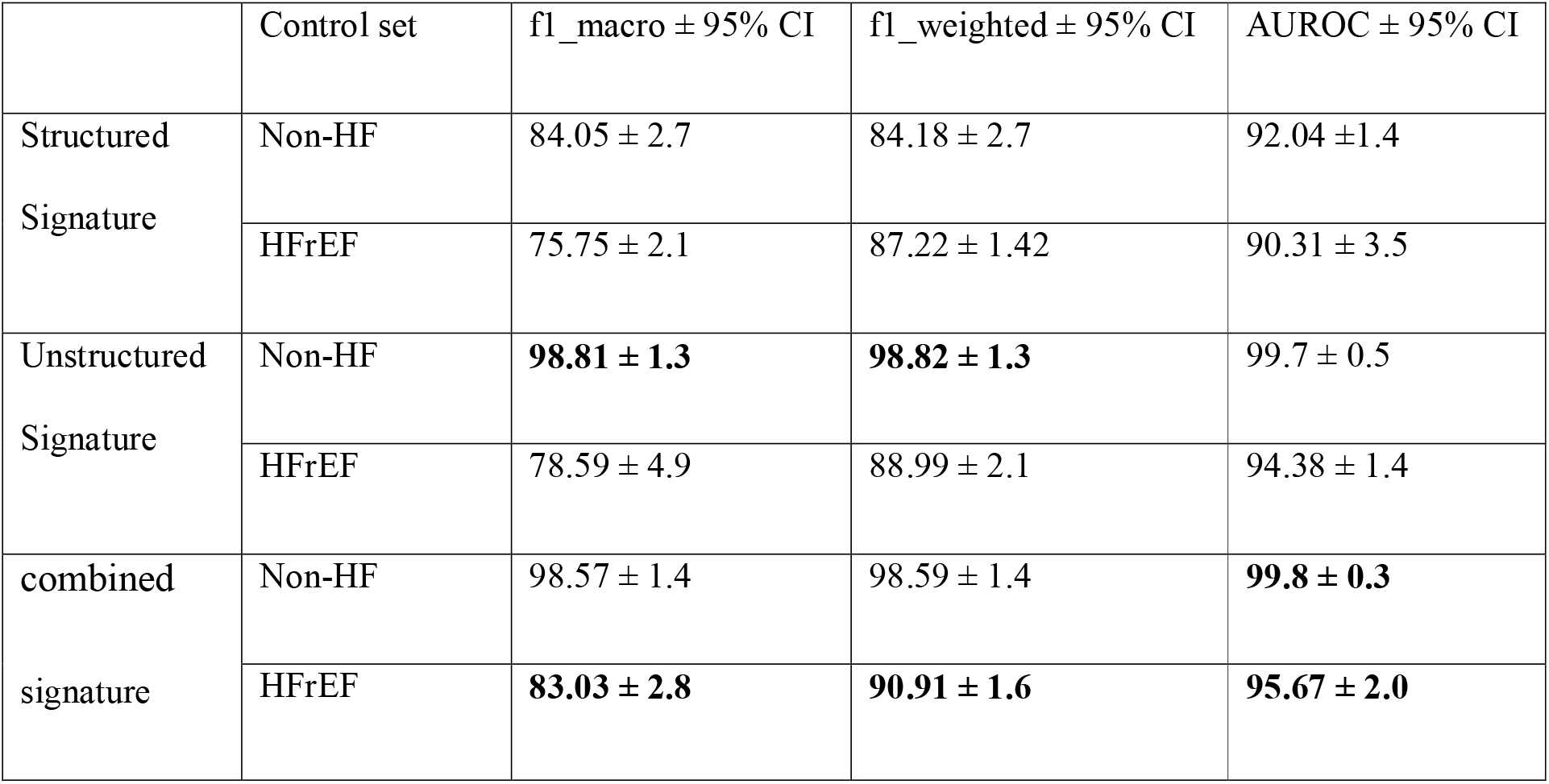
Multivariable model performance using the 5-fold cross-validation in derivation dataset.

**Table 3.**
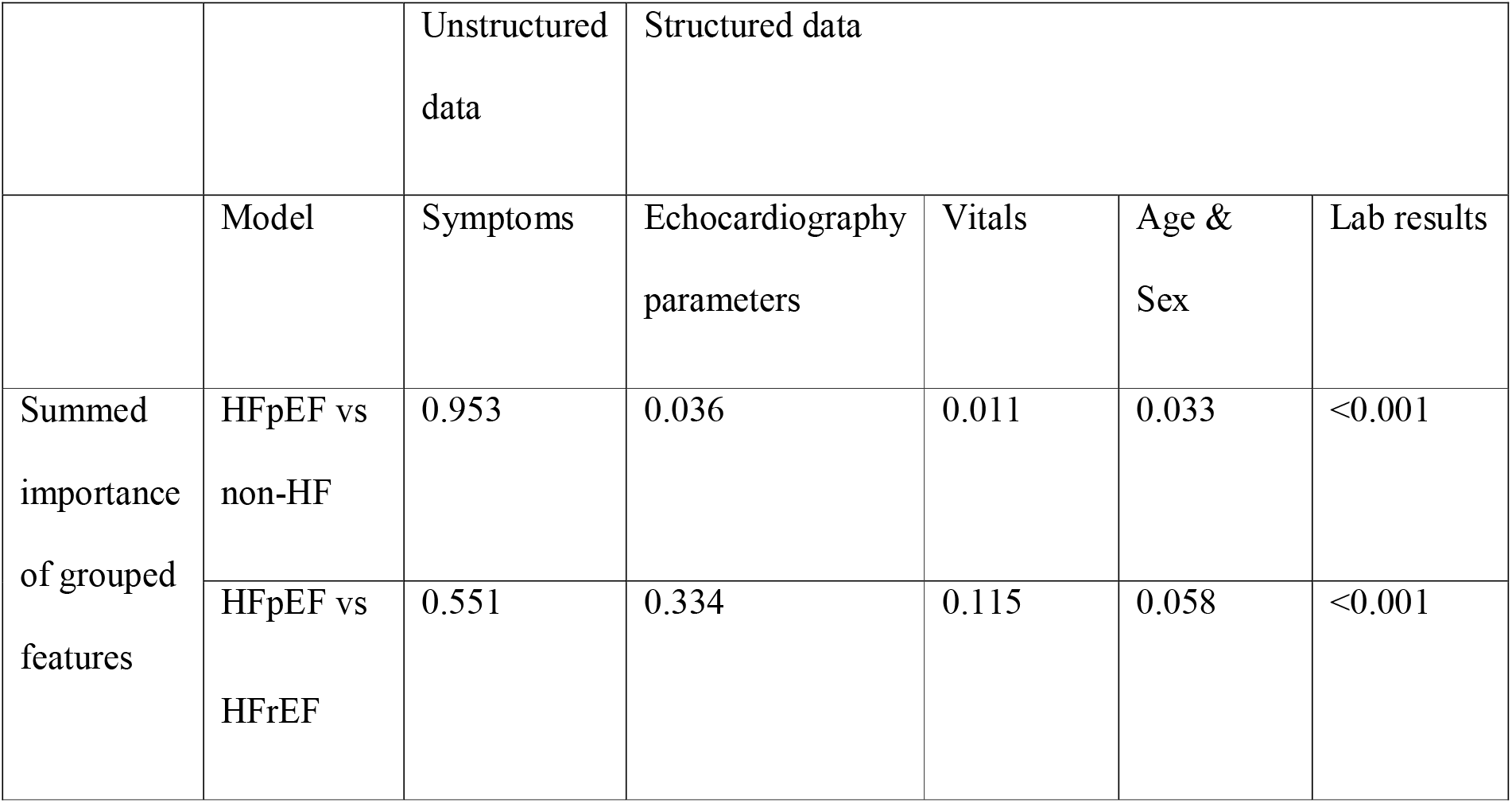
Additive SHAP feature importance for each category of predictors in the combined signatures.

### Selection of the final model and evaluation in test cohorts

The final model that was used for test evaluations aggregates the HFpEF vs HFrEF and HFpEF vs non-HF signature likelihood predictions, through an averaging operation. We used this aggregate model to make predictions on the test sets. **Figure S5** summarises the entire processing and model training pipeline.

To address the distributional variation between training and test cohorts which was caused by sample selection bias, we used 30% of the test samples (test1: 19, test2: 21, test3: 17, test4: 23) to retune the models, following the domain adaptation transfer learning technique[32]. Details of the 30% choice of adaptation set size is included in the **Supplementary materials, Figure S6**.

The performance of both proposed base models and the final aggregated model remained robust in the test cohort as compared to expert clinical consensus, with an AUROC performance of 0.86 (95% CI, ± 0.002) and 0.85 (95% CI, ± 0.001) in HFpEF vs non-HF and HFpEF vs HFrEF models, respectively and an enhanced aggregate performance of 0.90 (95% CI, ± 0.002) in our final aggregate model (**Figure 3**).

**Figure 3.**
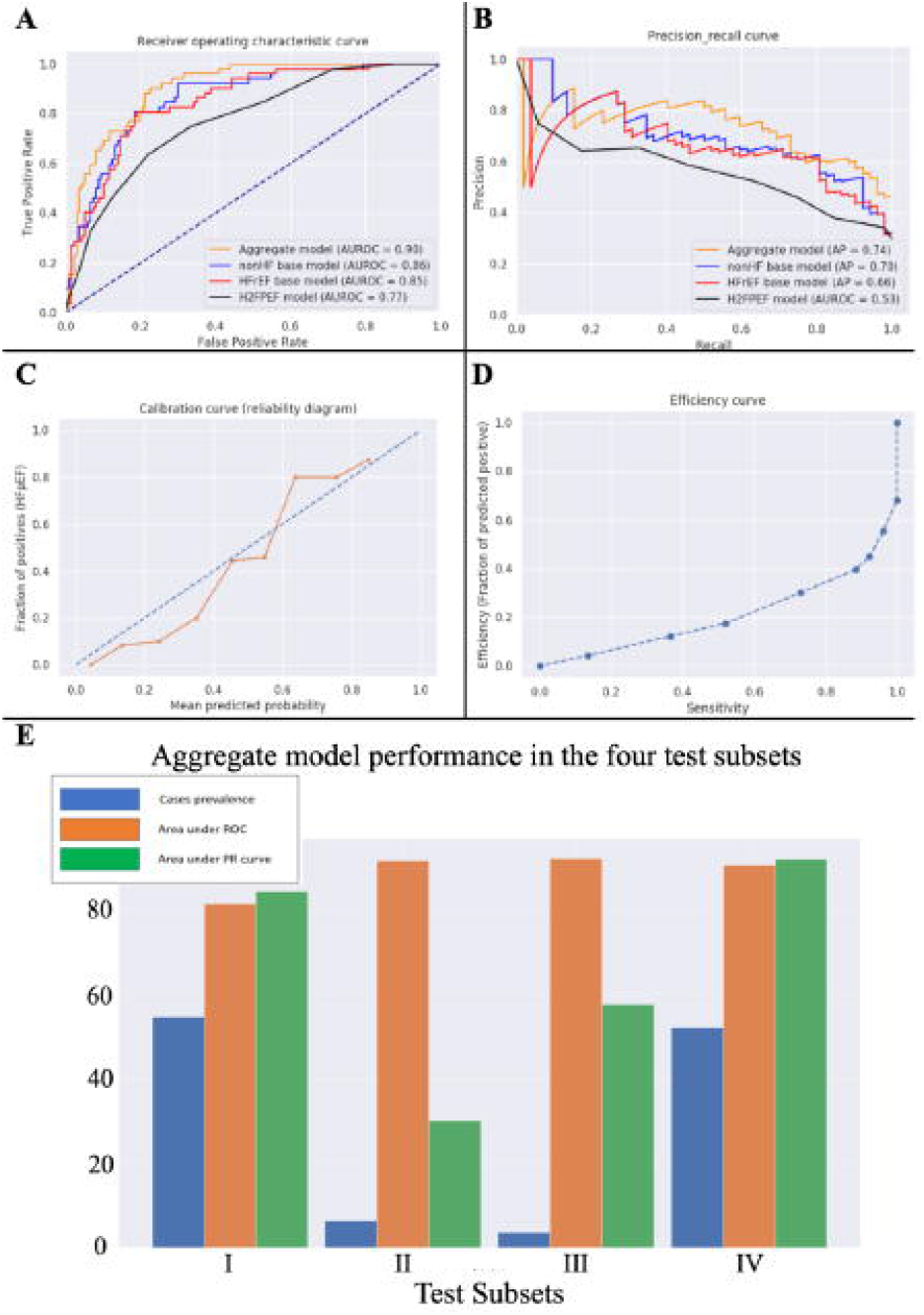
Performance of base and aggregate models. Panel A: Receiver Operating Characteristic curves for base models, aggregate model, and H_2_FPEF score. Panel B: Precision Recall curves for base models, aggregate model, and H_2_FPEF score. Panel C: Calibration curve for aggregate model. Panel D: Efficiency curve for aggregate model. Panel E: Aggregate model performance in the 4 test subsets

Lastly, we compared the final aggregate model as well as the baseline combined signature models (discriminating against non-HF or HFrEF) with the recently described H_2_FPEF model[16]. The AUROC and average precision of both the aggregate model and the individual baseline models was higher than the H_2_FPEF model (**Table 4**). We additionally used the Cohen’s kappa score to report on the agreement between the predictions made by our proposed models to better highlight the efficiency of the aggregate model over the individual base models discriminating HFpEF from non-HF and HFrEF. The positive kappa score of 0.3 indicates a weak agreement between the two base models. This was expected as the test cohort had lower availability of clinical assessments compared to the derivation cohort.

**Table 4.**
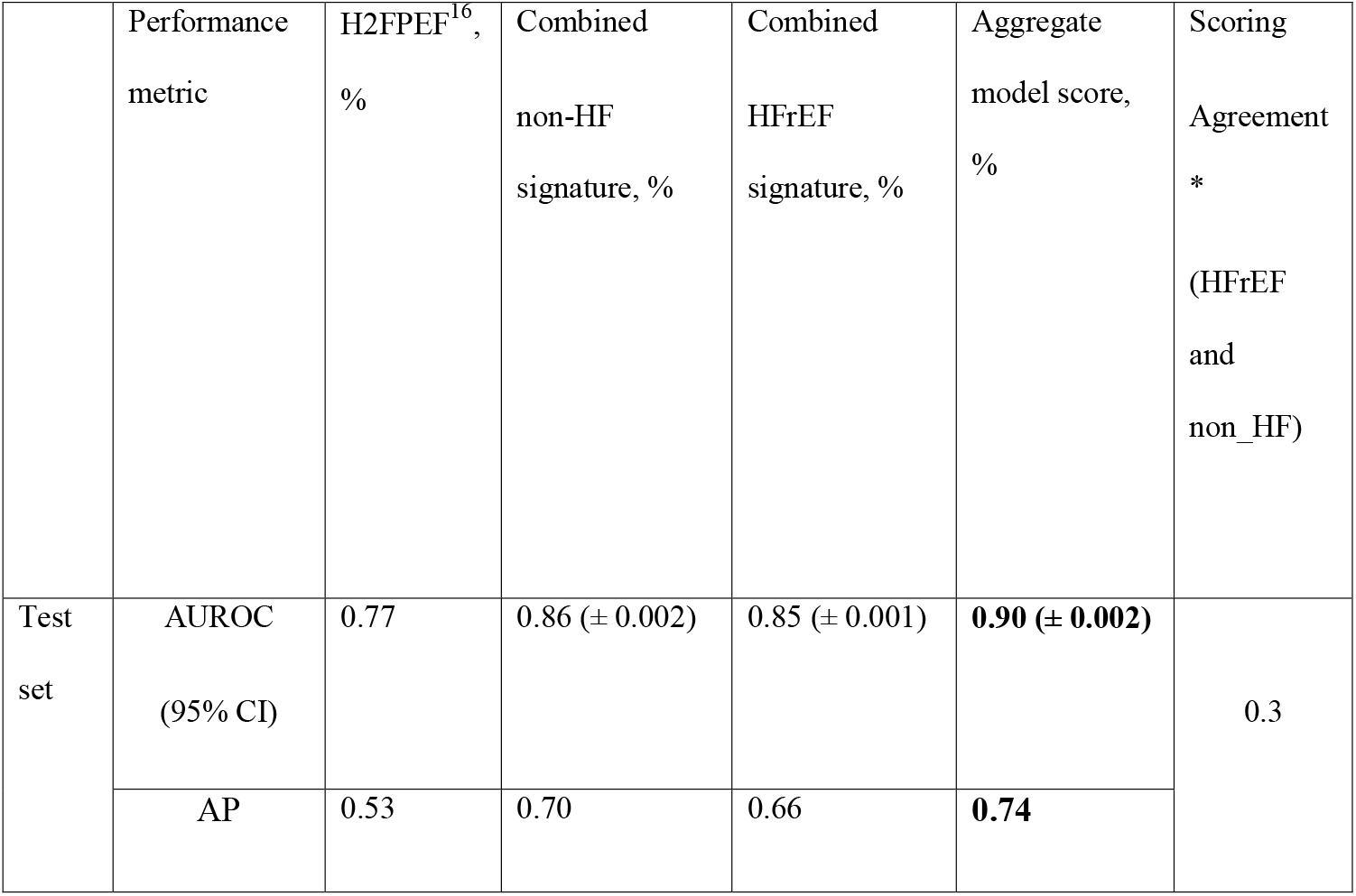
Multivariable model performance in independent test cohort. The 95% CI is reported using bootstrapping in a thousand of iterations. *: HFpEF annotation agreement between the two scoring systems using Cohen’s kappa statistics (python 3, Sklearn v.0.22). AUROC: area under receiver operative curve, AP: average precision, CI: confidence interval in bootstrapped samples

## DISCUSSION

In this study, we have developed an automated pipeline for EHR-based data collection, processing and modeling to identify patients with a high likelihood of HFpEF. We incorporated multi-modality data, including both structured and unstructured predictors, to generate a disease diagnostic signature. The proposed signature was validated in a separate cohort of patients and performed favourably as compared either to expert clinical consensus or the recently proposed H_2_FPEF score[16].

Analysis of the signatures that distinguished HFpEF from non-cardiac causes of dyspnea (non-HF) revealed anticipated predictors such as atrial fibrillation, hypertension, diabetes mellitus, kidney failure and obesity, in accordance with previous literature[16]. In addition, surrogate measures of multiple previous clinical encounters detected by the NLP algorithm as frequent text references to terms such as “pharmacologic substance” or “patient address” were very useful.

This may reflect the fact that patients with HFpEF may require multiple clinical visits and investigations, often with different specialities, before a diagnosis is established[17]. Apart from LVEF itself, features that distinguished HFpEF from HFrEF included age, peripheral edema, and other echocardiographic measures. An advantage of the approach that we employed may be that it is unbiased and comprehensive and identifies variables for inclusion in the diagnostic signature based purely on the results of the objective feature selection process. This may be one reason why our algorithm outperforms the H_2_FPEF score, which is based on the evaluation of selected variables rather than a comprehensive unbiased analysis. In this regard, it is of interest that echocardiographic predictors that contributed to the differentiation of HFpEF from HFrEF included maximum flow velocity across the aortic valve, aortic insufficiency and LA volume whereas E/e’ (which is part of the H_2_FPEF score) did not feature in the top 30 predictors.

A major underlying problem in efforts to develop or test new treatments for HFpEF is the difficulty in consistently diagnosing the syndrome[17]. Many different approaches are used in the literature based on varying criteria published by national and international societies, and diverse inclusion criteria have been used in clinical trials[33-35]. The problem is compounded by the likelihood that HFpEF is a heterogenous syndrome in which sub-populations may have differing underlying pathophysiology and outcomes[14, 15, 33]. The approach we present enables rapid identification of likely HFpEF cases among which further specific phenotyping could be performed to refine the diagnosis and potentially test or target defined interventions, or to identify potential subjects for research studies. Importantly, this approach aims to identify both compensated and decompensated HFpEF cases, using an automated and data-driven approach that is effective even where structured data (e.g. NT-proBNP measurements) are scarce. The approach may be considered complementary to scores such as H_2_FPEF. Our signature is ideally suited to rapidly identify a large number of possible HFpEF cases from EHR whereas H2PEF is better suited for use by the clinician evaluating an individual patient who is suspected to have HFpEF.

This study is the first to use SHAP analysis for feature selection in this context. We comprehensively validated all variations of the derived models in multiple datasets with underlying variational distributions. We demonstrated a significant improvement in HFpEF diagnostic performance when discriminating the patients with HFpEF from those with HFrEF or no HF history. A key strength of our approach is that modeling numerical assessment data (structured results signature) and EHR concept references separately makes the models applicable in scenarios where one of these sources of data may be scarce. Moreover, the dual modeling of HFpEF separation from non-HF and HFrEF subjects increases the utility of the proposed pipeline in distinguishing among a wider group of clinical conditions.

### Limitations

The UMLS clinical concept encoding that was used to extract unstructured observations does not support distinct encoding of different disease stages and could therefore cause some inaccuracy. In a more general aspect, the *a priori* assumptions that we made to identify definite HFpEF cases in the derivation dataset influenced the characterisation of the cohort. For example, we utilised ICD-10 diagnostic codes in the identification of patients with heart failure. Previous studies have demonstrated inaccuracy in identifying incident heart failure using ICD-10 coding as the sole source[36]. It is possible that such inaccuracy is present in our coding system; however the use of additional features (symptoms, LVEF, BNP/NTproBNP) in case classification mitigates this risk in our study. The inclusion of a raised BNP criterion restricts the cohort to a subgroup of HFpEF subjects, which was evident in test cohorts where many of the subjects did not have BNP measurements. This issue could be successfully handled through transfer learning techniques but would require some labelled data from a new domain to facilitate such a feedback training loop. The choice of data imputation technique could be another source of minor but systematic error. The discriminant power of the model to detect HFpEF is lower in test subsets where the missing data rate is higher and HFpEF cases are a small proportion of the overall number. Finally, the applicability of our model in patients with HFpEF who have never required hospital evaluation or admission is unknown. However, a strength of our approach is that a dedicated specialist assessment for HF is not required to assess the probability of HFpEF among patients undergoing general hospital evaluation (e.g. non-cardiological), even in the absence of commonly used diagnostic data such as NTproBNP levels. The lack of independent validation is a limitation of this study. Evaluation of the derived model’s performance in independent datasets from other centres and in community-based datasets will be informative in future studies. Although we compared performance of the model with the H_2_FPEF score,[16] due to its stated aim of estimating the likelihood that HFpEF among patients with unexplained dyspnoea to guide further testing, we did not compare performance to the HFA-PEFF algorithm[37] which is a multi-step diagnostic algorithm. Furthermore, the comparison of our alogrithm’s performance with the H_2_FPEF should be confirmed in a separate validation cohort.

## Conclusion

In this study, we have developed a rapid and automated data-driven approach that is effective at identifying patients from EHR who are likely to have HFpEF. This algorithm affords significant potential to rapidly identify patients for more detailed analyses and/or potential inclusion in clinical trials. The approach that we report could in principle be readily applied to other diseases and conditions that are similarly difficult to diagnose.

## Supplemental Materials

The supplementary digital content is provided to support the findings of this study.

## Contributors

Study design: NF, KO, RD, AMS;

Data collection: NF, JK, JO, AM;

Data modeling: NF, DB, RD;

Data analysis: NF, KO;

Clinical validation: KO, RZ, DB2, AN;

Result interpretation and writing the paper: All authors.

Funding: AMS.

Supervision: RD and AMS.

## Acknowledgements

We thank Ahmed Mahmmud and Joe Omigie for their invaluable advice during the data collection phase and Norman Catibog and Thiago Fonseca for sharing their knowledge in echocardiography. This work was supported by the British Heart Foundation (RE/18/2/34213; CH/1999001/11735); the NIHR Biomedical Research Centres at Guy’s & St Thomas’ NHS Foundation Trust (IS-BRC-1215-20006) and South London and Maudsley NHS Foundation Trust (IS-BRC-1215-20018), both with King’s College London. KOG is supported by a Medical Research Council Clinical Training Fellowship (MR/R017751/1). DMB is funded by a UKRI Innovation Fellowship as part of Health Data Research UK MR/S00310X/1 (https://www.hdruk.ac.uk). The views expressed are those of the authors and not necessarily those of NIHR or the Department of Health and Social Care. The funders had no role in study design, data collection and analysis, decision to publish, or preparation of the manuscript.

## Data Sharing

The raw data used in this research are not openly available.

