## Supplementary Material for "Diagnostic signature for Heart Failure with Preserved Ejection Fraction (HFpEF): A Machine Learning Approach Using Multi-Modality Electronic Health Record Data"

### Sec. I Diagnosis assertion criteria

***Table S1.*** *Data availability table used for case and control definition and cohort selection. Black shaded cells indicate the validity of a criterion for a set, grey shade indicates partial validity and the white indicates criterion in a given set with an exception on n/rBNP for test set I, II and III where no BNP result was available for these subsets. HF: heart failure, Diag: ICD10 diagnosis, cDiag: complementary diagnosis derived from semi-structured data through NLP, pEF: preserved EF (>=50), rEF: reduced EF (<50), nBNP: normal BNP, rBNP: abnormal BNP*

| Sets | HF concept ref. | Diag | cDiag | pEF | rEF | nBNP | rBNP | Dysp |
|---|---|---|---|---|---|---|---|---|
| **DERIVATION COHORT** | | | | | | | | |
| Confirmed HFpEF | | | | | | | | |
| Control I (Non-HF) | | | | | | | | |
| Control II (HFrEF) | | | | | | | | |
| **TEST COHORT** | | | | | | | | |
| Test subset I | | | | | | | | |
| Test subset II | | | | | | | | |
| Test subset III | | | | | | | | |
| Test subset IV | | | | | | | | |

***Flowchart S1.*** *The cohort definition flowchart*

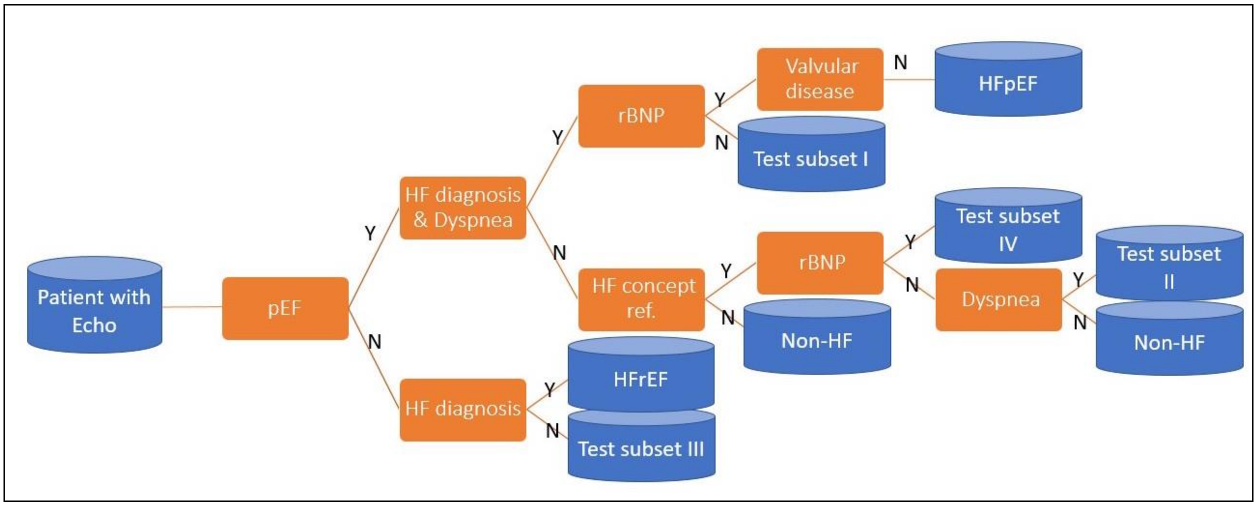

***Figure S1.*** *Age and gender histogram in the HFpEF cases of derivation set.*

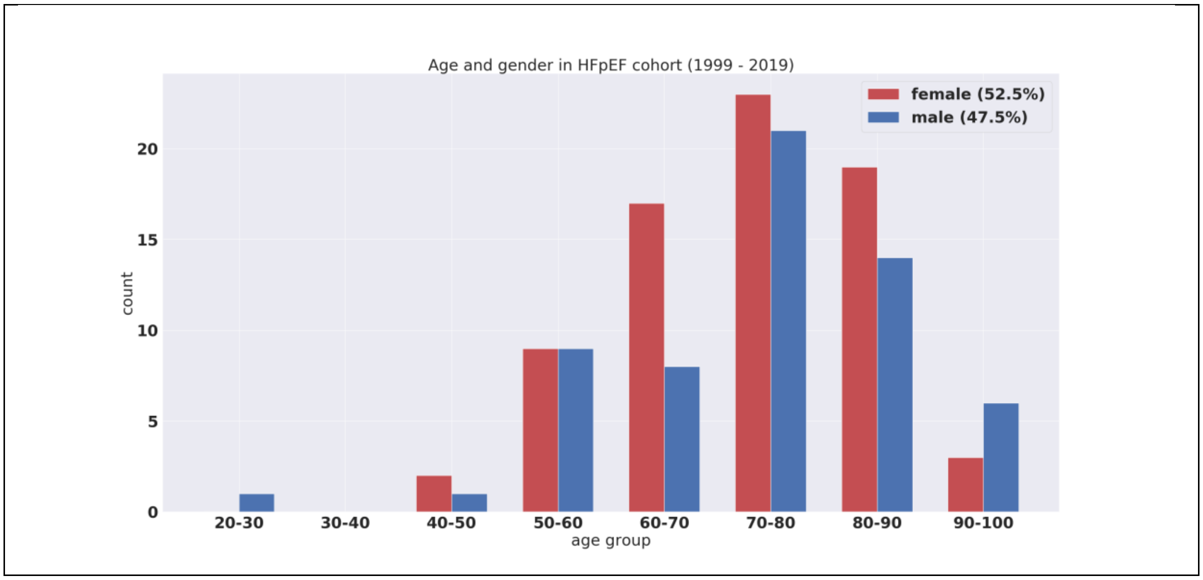

### Sec. II: BNP/NT-proBNP assessment criteria

***Alg.1.*** *Algorithm which was used to define the HFpEF cases for the study*

```
Inputs: TEST_TYPE, BNP_VALUE, PATIENT_AGE
Output: BNP_state

If TEST_TYPE is NT-proBNP:
        If (BNP_VALUE >= 125) & (PATIENT_AGE <75):
                BNP_state <-- abnormal
        Else if (BNP_VALUE>=450) & (PATIENT_AGE >=75):
                BNP_state <-- abnormal
        Else:
                BNP_state <-- normal
If TEST_TYPE is BNP:
        If (BNP_VALUE >= 100) & (PATIENT_AGE <75):
                BNP_state <-- abnormal
        Else if (BNP_VALUE>=130) & (PATIENT_AGE >=75):
                BNP_state <-- abnormal
        Else:
                BNP_state <-- normal
Return BNP_state
```

**Sec. III: Ejection fraction aggregation across structured and unstructured data**

In order to extract all EF values in patient's historical records, we used two sources of data; 1) structured data containing echocardiographic measurements of in-hospital Ultrasounds scans and 2) unstructured free-text reports of the ultrasound results obtained in other hospitals and included in patient's EPR.

Extracting the EF values from structured data is done through SQL queries. The EF info in the text, however encompassed in three main formats, i) numeric EF reported within the text, ii) semi-structured fields inserted as text in the EPR document and iii) the clinical interpretation on normality of the EF value is reported in free-text (Figure S2 depicts these variation).

To cope with all variations, we used three parallel streams of natural language processing units. The first processing units (PU) extracts the numeric reports of the EF measurements in a word-window around the EF reference in the text using NLTK python toolbox. The second unit uses a rule-based approach to re-structure the semi-structured echocardiographic reports into a SQL table. The numerical results of the first two units are aggregated with the structured EF values on subject level.

For subjects who had no numerical EF values extracted but had echocardiographic interpretation summary, we used a three-layers LSTM network to infer the EF deviation from normality (positive class: EF>=50%, negative class: EF<50%).

We have tested the performance of the processing units on the mutually available reports in both structured and unstructured data sources.

The performance of our proposed deep learning approach is compared (Figure S3) with a decision tree model using the three-fold cross validation of ROC curves and on 2000 manually annotated samples. Finally, the staged aggregation evaluations are reported in Figure S4 using the subset of subjects who had EF assessment data availability across the two structured (Xcelera, SQL) and unstructured (EPR free-text, ELK) databases.

***Figure S2.*** *Various formats in which the EF value is reported in unstructured EPR reports. EF: Ejection fraction*

```
Interpretation Summary

A two-dimensional transthoracic echocardiogram with M-mode and Doppler was performed.
Normal biventricular size and systolic function. Other chambers and all valves are structurally normal.
Right ventricular systolic pressure is estimated at 40-45mmHg.

Left Ventricle
The left ventricle is normal in size. There is normal left ventricular wall thickness. Left ventricular
systolic function is normal. Estimated LVEF ~64% by Simpson's bi-plane method.
Abnormal Diastolic Function Grade I - Impaired Relaxation with normal filling pressures.
The left ventricular wall motion is normal....

Measurments

LVIDd: 3.9 cm (3.9-5.3 cm)

LVIDs: 2.5 cm (2.5-4.5 cm)

FS: 35.4 % (27-45%)

IVSd: 0.92 cm (0.8-1.1 cm)

LVPWd: 0.71 cm (0.5-1.1 cm)

LA dimension: 3.0 cm (2.7-3.8 cm)

DV(MOD-sp2): 105.9 ml

EDV(sp2-el): 102.4 ml

LVLs ap2: 6.6 cm

EF(MOD-sp2): 66.2 %

EF(sp2-el): 66.5 %

EF(MOD-bp): 64.3 %
...
```

***Figure S3.*** *Deep learning model performance on discriminating the text reference of a preserved/normal ejection fraction from the reduced fraction.*

| Transductive LSTM | Decision Tree |
| --- | --- |

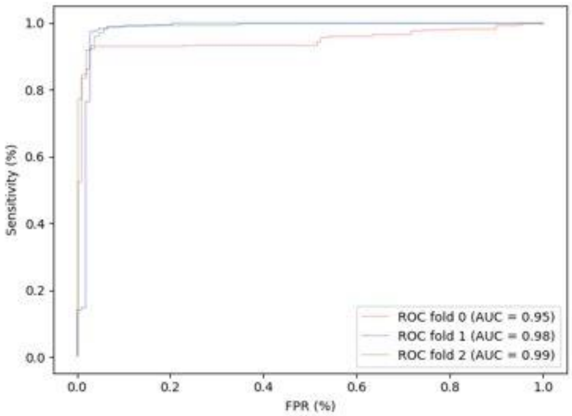


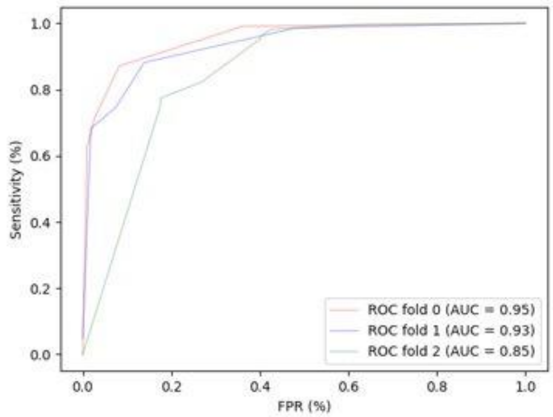


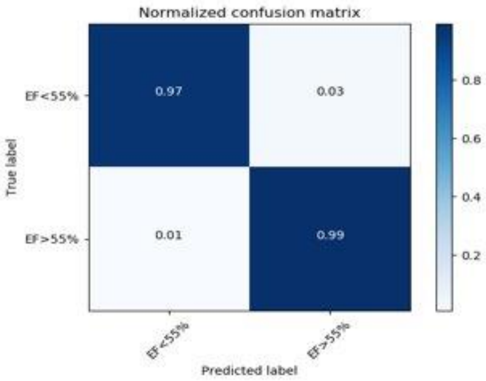


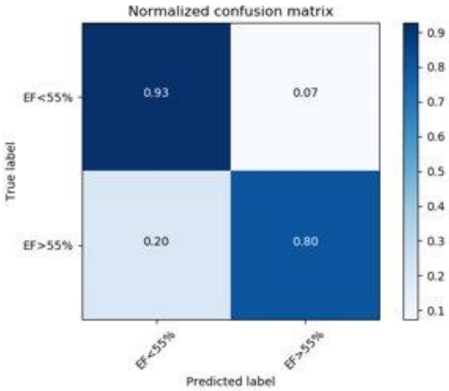

***Figure S4.*** *NLP pipeline validation in subjects with EF data availability across structured and unstructured platforms. The aggregation was aimed at obtaining a comprehensive history of all ejection fraction assessments performed for a patient throughout his medical history.*

<table>
<tr><td>Deep learning model validation</td><td>Overall text extracted EF aggregation validation</td></tr>
<tr><td>
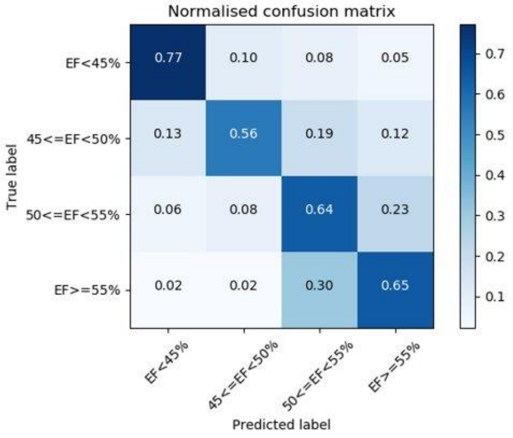

</td><td>
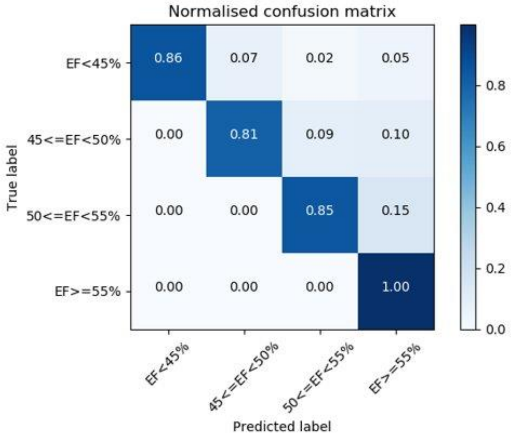

</td></tr>
</table>

### Sec. IV: Data processing and signature training pipeline

***Figure S5.*** *Data processing and machine learning pipeline for automated diagnosis of patients suffering from HFpEF of any disease stage.*

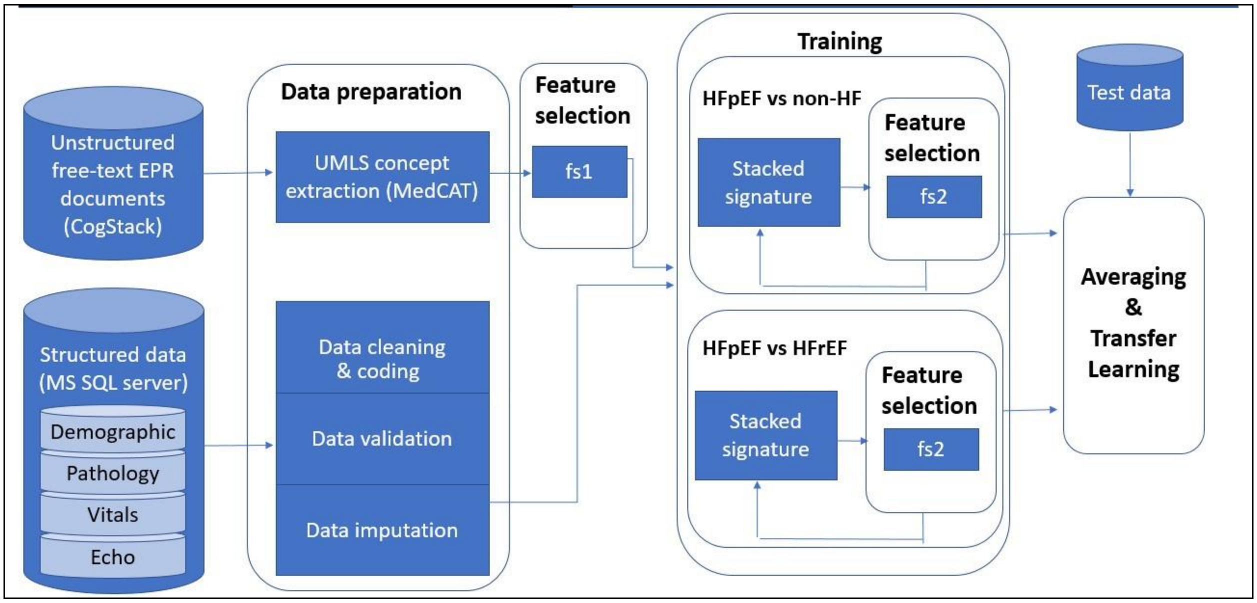

### Sec. V: Echocardiographic measurements performed in KCH

***Table S2.*** *Echocardiographic measurements and their unit of assessment used in the study. LV: Left ventricle, RV: Right ventricle, MV: Mitral valve, TV: Tricuspid valve, AV: aortic valve, HM: Heart model, BSA: Body surface are, PK EX: Peak exercise.*

| LABEL | Unit of measurement | DESCRIPTION |
|---|---|---|
| AI MAX VEL | cm/sec | Aortic valve insufficiency max velocity |
| AO LVETC | sec | Aortic outflow (doesn't exist!!!) |
| AO MEAN (PK EX) | cm/sec | Aortic outflow peak velocity at peak exercise |
| AO PEAK VEL (PK EX) | cm/sec | Aortic outflow mean velocity at peak exercise |
| AO V2 MAX | cm/sec | Aortic valve max velocity envelope of the distal flow |
| AO V2 MEAN | cm/sec | Mean velocity envelope of the distal flow in Aortic valve |
| AS MAX VEL | cm/sec | AS jet velocity max |
| AV PEAK VEL | cm/sec | Aortic valve peak velocity |
| AV VMAX-PR_PHL | cm/sec | Aortic valve systolic velocity prosthetic antegrade flow |
| E/E' AVERAGE | - | Mitral peak velocity of early filling (E) to early diastolic mitral annular velocity (E'), Average |
| E/E'_LAT | - | Mitral peak velocity of early filling (E) to early diastolic mitral annular velocity (E'), Lateral |
| E/E'_SEPT | - | Mitral peak velocity of early filling (E) to early diastolic mitral annular velocity (E'), Septal |
| EF (manual) | % | Ejection fraction |
| LVEF (automated HM) | % | Ejection fraction (heart model, automated) |
| IVR TIME | sec | Isovolumic relaxation time |
| LA AREA | cm^2 | Left atrial area |
| LA DIMENSION | cm | Left atrial dimension |
| LA SYSTOLIC VOLUME (HM) | ml | Left atrial systolic volume |
| LA VOL (2D BIPLANE) INDEX BSA | ml/m^2 | Left atrial volume normalised to body surface area |
| LA VOLUME (2D BIPLANE) | ml | Left atrial volume (2d biplane) |
| LA/AO | - | LA/AO |
| LV DP/DT | mmHg/s | Difference in the pressure through mitral valve inflow in time (isovolumic phase index of the LV systolic performance) |
| LV EDV (HM) | ml | LV end diastolic vol |
| LV ESV (HM) | ml | LV end systolic Vol |
| LV EXCURSION - AVG | cm | Mitral annular plane systolic excursion (MAPSE), average |
| LV EXCURSION - MAX | cm | Mitral annular plane systolic excursion (MAPSE), max |
| LV EXCURSION - MIN | cm | Mitral annular plane systolic excursion (MAPSE), min |
| LV EXCURSION - SD | cm | Mitral annular plane systolic excursion (MAPSE), std |

| | | |
|---|---|---|
| LV MASS - END DIAS | g | LV mass - end diastolic |
| LV MASS - END SYS | g | LV mass - end systolic |
| LV MASS(AL) DIAS INDEXED | g/m^2 | LV mass |
| LV MASS(C) SYS INDEXED | g/m^2 | LV mass |
| LV PK 1 | cm/sec | Max velocity of the flow proximal to the AV at stress echo (rest) |
| LV PK 2 | cm/sec | Max velocity of the flow proximal to the AV at stress echo (low dose) |
| LV PK 3 | cm/sec | Max velocity of the flow proximal to the AV at stress echo (peak) |
| LV STROKE VOLUME (HM) | ml | LV stroke volume |
| LV V1 MAX | cm/sec | Left ventricular max velocity proximal to AV |
| LV V1 MAX (PK EX) | mmHg | Left ventricular max pressure gradient at peak exercise |
| LV V1 MAX PG | mmHg | Left ventricular max pressure gradient |
| LV V1 MEAN | cm/sec | Left ventricular outflow tract mean velocity |
| LV V1 MEAN PG | mmHg | Left ventricular outflow tract mean velocity |
| LV V1 MEAN PG (PK EX) | mmHg | Left ventricular outflow tract mean velocity at peak exercise |
| LV V1 VTI | cm | Left ventricular outflow tract velocity time integral |
| LV V1 VTI (PK EX) | cm | Left ventricular outflow tract velocity time integral |
| LV VMAX 10MCG | mmHg | Max pressure gradient of flow proximal to AV |
| LV VMAX REST | mmHg | Max pressure gradient of flow proximal to AV at rest |
| LV VOL D (INDEXED BSA) | ml/m^2 | LV volume diastolic normalised to body surface area |
| LV VOL S (INDEXED BSA) | ml/m^2 | LV volume systolic normalised to body surface area |
| LV VTI 10MCG | cm | Left ventricular outflow tract velocity time integral |
| LV VTI 5MCG | cm | Left ventricular outflow tract velocity time integral |
| LV VTI REST | cm | Left ventricular outflow tract velocity time integral at rest |
| LVAD SAX MAX | cm^2 | LV area at diastolic |
| LVAS SAX MV | cm^2 | LV area at systole |
| LVD MASS/VOLUME | - | LV diastolic mass to volume |
| LVDIAS VOL 3D (INDEXED BSA) | ml/m^2 | LV diastolic vol 3D |
| LVET | sec | Left ventricle Ejection time |
| LVIDD | cm | Left ventricular internal diameter end diastole |
| LVIDD (INDEXED BSA: CM/M^2) | - | Left ventricular internal diameter end diastole (normalised to BSA) |
| LVIDS | cm | Left ventricular internal diameter end systole |
| LVLD %DIFF | % | LV long axis difference ration at diastole |
| LVLD APICAL(4CH) | cm | LV long axis apical contour at diastole (4-chamber view) |
| LVLD EPI APICAL(4CH) | cm | LV epicardial long axis apical contour at diastole (4-chamber view) |
| LVLS %DIFF | % | LV long axis difference ration at systole |
| LVLS AP2 | cm | LV long axis at systole (2-chamber view) |
| LVLS AP4 | cm | LV long axis at systole (4-chamber view) |

| | | |
|---|---|---|
| LVLS APICAL | cm | LV long axis apical contour at diastole (4-chamber view) |
| LVOT ACCEL SLOPE | cm/sec^2 | LV outflow tract ACCEL slop (cm/s^2) |
| LVOT _AREA | cm^2 | LV outflow tract area (automated possibly) |
| LVOT AREA | cm^2 | LV outflow tract area (automated) |
| LVOT AREA(TRACED) | cm^2 | LV outflow tract area (manual more accurate) |
| LVOT D (2D) | cm | LV outflow tract diameter |
| LVOT DIAM | cm | LV outflow tract diameter |
| LVPEP | sec | LV pre-ejection period |
| LVPWD | cm | Left ventricular posterior wall end diastole |
| LVPWS | cm | Left ventricular posterior wall end systole |
| LVS MAJOR | cm | LV major dimension at diastole |
| LVSYS VOL 3D (INDEXED BSA) | ml/m^2 | LV systolic vol 3D indexed to BSA |
| MAX V | cm/sec | Max doppler velocity |
| MEAN V | cm/sec | Mean doppler velocity |
| MED PEAK S VEL | cm/sec | Peak s-wave velocity of the medial MV annulus tissue doppler spectrum |
| MITRAL R-R | sec | Mitral R-R time |
| MM R-R INT | sec | M-mode R-R interval |
| MR ALIAS VEL | cm/sec | Mitral regurgitation alias velocity |
| MR MAX VEL | cm/sec | Mitral regurgitation max velocity |
| MR MEAN VEL | cm/sec | Mitral regurgitation mean velocity |
| MV A DUR | sec | Mitral valve A-wave duration |
| MV A MAX VEL | cm/sec | Mitral valve A-wave max velocity |
| MV A MAX VEL (PK EX) | cm/sec | Mitral valve A-wave max velocity (peak exercise) |
| MV ACC TIME | sec | Mechanical ventilation acceleration time (MV: Mitral valve) |
| MV DEC SLOPE (PK EX) | cm/sec^2 | Mechanical ventilation deceleration slope (MV: Mitral valve) |
| MV DEC TIME | sec | Mechanical ventilation deceleration time (MV: Mitral valve) |
| MV DEC TIME (PK EX) | sec | Mechanical ventilation deceleration time (peak exercise) |
| MV E MAX VEL | cm/sec | Mitral valve E-wave max velocity |
| MV E/A | - | Mitral valve E-wave/A-wave ratio |
| MV E-F SLOPE | cm/sec | Mitral valve E-F slope |
| MV P1/2T MAX VEL | cm/sec | Mitral valve P1/2T max velocity |
| MV V2 MAX | cm/sec | Mitral valve V2 (highest velocity at valve) max |
| MV V2 MAX (PK EX) | cm/sec | Mitral valve V2 (highest velocity at valve) max (peak exercise) |
| MV V2 MEAN | cm/sec | Mitral valve V2 (highest velocity at valve) mean (MV: Mitral valve) |
| PA ACC TIME | sec | Pulmonary velocity acceleration time |
| PA ACCEL TIME | sec | Pulmonary velocity acceleration time |
| PA V2 MAX | cm/sec | Pulmonary velocity max |
| PA V2 MEAN | cm/sec | Pulmonary velocity mean |
| PI MAX VEL | cm/sec | Pulmonary insufficiency max velocity |

| | | |
|---|---|---|
| RAP systole | mmHg | Right atrial pressure at systole |
| RAP diastole | mmHg | Right atrial pressure at diastole |
| R-R TIME | sec | R-R time |
| RV V1 MAX | cm/sec | Right ventricle outflow tract max velocity |
| RV V1 MEAN | cm/sec | Right ventricle outflow tract mean velocity |
| TR MAX PG | mmHg | Tricuspid regurgitation max pressure gradient |
| TR MAX PG (PK EX) | mmHg | Tricuspid regurgitation max pressure gradient (peak exercise) |
| TR MAX VEL | cm/sec | Tricuspid regurgitation max velocity |
| TR MAX VEL (PK EX) | cm/sec | Tricuspid regurgitation max velocity (peak exercise) |
| TR VTI | cm | Tricuspid regurgitation velocity time integral |
| TV A MAX VEL | cm/sec | Tricuspid valve A-wave max velocity |
| TV DEC TIME | sec | Tricuspid valve deceleration time |
| TV E MAX VEL | cm/sec | Tricuspid valve E-wave max velocity |
| TV E/A | - | Tricuspid valve E/A ratio |
| TV P1/2T MAX VEL | cm/sec | Tricuspid valve P1/2T max velocity |
| TV V2 MAX | cm/sec | Tricuspid valve V2 max |
| TV V2 MEAN | cm/sec | Tricuspid valve V2 mean |
| VSD MAX VEL | cm/sec | Ventricular septal defect max velocity |

**Sec. VI: Performance metrics**

F1-measure score = 2*precision*recall / (precision + recall)

Kappa score = (observed agreement – expected agreement) / (1 - expected agreement)

Expected agreement using the observed data is defined as the probabilities of each classifier model randomly predicting each category. This in effect for our binary classification task translates into:

Expected agreement = (n11*n12 + n21*n22)/N^2

Where N is the total number of samples (=269) and nik indicates the number of times the classifier i predicted category k (k in our study can take either of values, 0 indicative of no HFpEF and 1 indicative of HFpEF diagnosis.)

### Sec. VII: Model adaptation

***Figure S6.*** *Model adaptation optimization: different proportion of the test subject data were used to retune the model parameters for test-set usability and generalisation. Orange: AUROC performance metric, Blue: average precision (AP) performance metric.*

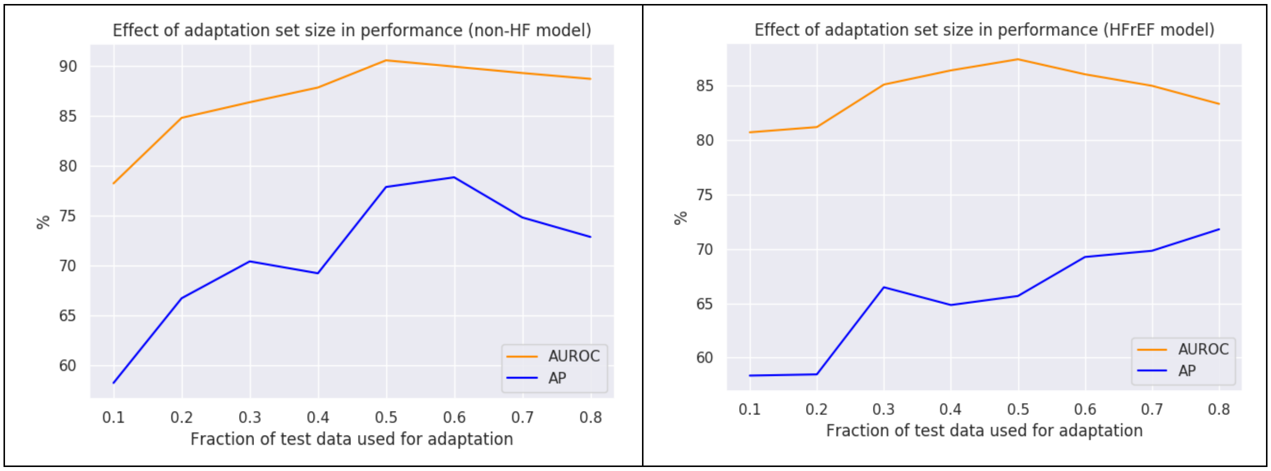

### Sec. VIII: H2FPEF score computation

To compute the H2FPEF we have automated the data query for the six parameters that contribute in the score estimation. Missing values of the BMI were estimated using the patient's weight and height (BMI = weight/(height)**2). We have used the average value of the e/e' septal and lateral for filling pressure and the sum over the maximum tricuspid regurgitation pressure gradient and the right atrial pressure at systole[22] (where available) as the closets estimate of the pulmonary artery systolic pressure, proposed by [16]. The ICD10 code of I48*** has been used for ascribing prior diagnosis of atrial fibrillation. And the data from the UMLS drug hierarchy was queried to verify whether a prescribed medication is an antihypertensive drug - we used simple regex text cleaning, the UMLS repository and MedCAT medical text annotation tool to accomplish this. Following the linkage of these six data items for each patient, missing values on each item were replaced with the normal value of that parameter - replacements with abnormal values resulted in lower AUROC value for H2FPEF score - i.e. missing values of BMI were replaced by 29 kg/m2.

Measurements for each data item were then ascribed with scoring points following[16]. Figure S7 summarizes the scoring system proposed by[16].

***Figure S7.*** *H2FPEF scoring system.*

| | Clinical Variable | Values | Points |
|---|---|---|---|
| $H_2$ | Heavy | Body mass index > 30 kg/m$^2$ | 2 |
| | Hypertensive | 2 or more antihypertensive medicines | 1 |
| F | Atrial Fibrillation | Paroxysmal or Persistent | 3 |
| P | Pulmonary Hypertension | Doppler Echocardiographic estimated Pulmonary Artery Systolic Pressure > 35 mmHg | 1 |
| E | Elder | Age > 60 years | 1 |
| F | Filling Pressure | Doppler Echocardiographic E/e' > 9 | 1 |
| $H_2$FPEF score | | | Sum (0-9) |

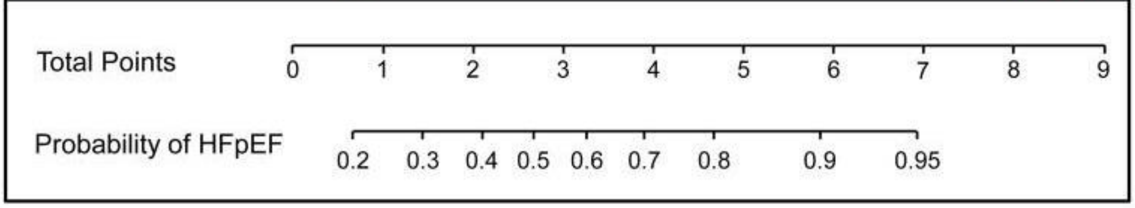

**Sec IX: FN/FP analysis**

***Table S3:*** *Aggregate model False negative and False positive assessments. The highlighted lines are indicative of the FP cases where our Aggregate model assigned a HFpEF label incorrectly.*

| Agg. Model Pred. | H2FPEF Score | Anno. | Clinical comments |
|---|---|---|---|
| 1 | 3 | 0 | Alternate diagnosis: COPD which explains the breathlessness. Preserved LV function, and mildly dilated LA and LVH, but no other heart failure symptoms apart from dyspnoea. |
| 0 | 8 | 1 | Does have a diagnosis of HFpEF |
| 0 | 2 | 1 | Does have a diagnosis of HFpEF - symptoms of heart failure, preserved EF on echo, positive BNP |
| 0 | 2 | 1 | Clinic letter from 2004 discussion diastolic dysfunction on echo, SOB and peripheral oedema. No BNP was ever reported. |
| 1 | 6 | 0 | Alternate diagnosis: AF causing pulmonary eodema, but this was precipitated by drugs in hospital |
| 1 | 8 | 0 | Two reports on the echocardiograms suggesting mildly or borderline reduced LV function |
| 1 | 4 | 0 | Alternate diagnosis: pulmonary emboli on a CT scan |
| 1 | 4 | 0 | Alternate diagnosis: SOB in context of pneumonia. Previous CABG for ischemic heart disease, but no consistent heart failure syndrome |
| 1 | 5 | 0 | Alternate diagnosis: end stage renal failure on dialysis. Episodes of fluid overload are not due to heart failure. Other episodes of breathlessness are in the context of infection. |
| 1 | 6 | 0 | Alternate diagnosis: severe aortic stenosis |
| 1 | 4 | 0 | Alternate diagnosis: breathlessness due to suspected infection |
| 1 | 4 | 0 | Alternate diagnosis: Breathlessness due to chest infection / reaction to antibiotic |
| 1 | 3 | 0 | Alternate diagnosis: Ischaemic heart disease |
| 1 | 3 | 0 | Alternate diagnosis: pulmonary hypertension due to pulmonary haemorrhage |
| 0 | 4 | 1 | Sickle cell patient. Does have some echo features of HFpEF but symptoms in the context of leukemia/pericardial effusion/presumed PE, therefore always an alternate diagnosis. |
| 1 | 5 | 0 | Alternate diagnosis: Severe tricuscpid regurgitation due to a pacemaker lead problem |
| 1 | 4 | 0 | Alternate diagnosis: diabetic patient with reduced EF on echo |
| 1 | 5 | 0 | Alternate diagnosis: Hypertrophic cardiomyopathy |
| 1 | 6 | 0 | Alternate diagnosis: Severe aortic stenosis |
| 1 | 9 | 0 | Alternate diagnosis: Right ventricular dysfunction |

| | | | |
|---|---|---|---|
| 1 | 2 | 0 | Alternate diagnosis: ischemic heart disease and moderate aortic regurgitation, lung disease causing breathlessness |
| 1 | 7 | 0 | Alternate diagnosis: Severe COPD with normal BNP |
| 1 | 4 | 0 | Alternate diagnosis: Aortic valve replacement |
| 1 | 6 | 0 | Alternate diagnosis: ischemic heart disease and pulmonary fibrosis |
| 1 | 4 | 0 | No consistent history of heart failure |
| 1 | 8 | 0 | Alternate diagnosis: Aortic valve disease |
| 1 | 8 | 0 | Alternate diagnosis: AF, Symptoms of oedema improved post-cardioversion |
| 1 | 3 | 0 | Alternate diagnosis: Severe lung disease |
| 1 | 4 | 0 | Alternate diagnosis: Right ventricular dysfunction |
| 1 | 8 | 0 | Alternate diagnosis: Ischaemic heart disease requiring multiple stents |
| 0 | 1 | 1 | Does have HFpEF Breathlessness, preserved LV function, positive BNP. On the initial echo in September 2019 the TR was only mild-moderate, so this is enough to make the diagnosis. Subsequently her TR worsened to severe |
| 1 | 6 | 0 | Alternate diagnosis -> pulmonary fibrosis |
| 1 | 5 | 0 | Alternate diagnosis: Hypertrophic cardiomyopathy |
